# Scaling ECG Foundation Models and Identifying a Threshold for Effective Representation Learning

**DOI:** 10.64898/2026.07.15.26358182

**Authors:** Raghav Sriram, Ivan Nenadic, Elan Shahrabani, Sascha N. Goonewardena, Shanshan Yao, Brennen Farrell, Zak Loring, Venkatesh L Murthy

**Affiliations:** Lampe Joint Department of Biomedical Engineering, North Carolina State University, Raleigh, NC; Division of Cardiology, Department of Medicine, Northwestern University, Feinberg School of Medicine Chicago, Illinois, United States; Duke Heart Center, Duke University Hospital, Durham, North Carolina, United States; Department of Internal Medicine, University of California in Los Angeles, Los Angeles, California, United States; Department of Internal Medicine, Division of Cardiovascular Medicine, University of Michigan, Ann Arbor, United States; Department of Computer Science, North Carolina State University, Raleigh, North Carolina, United States

## Abstract

We conducted a scaling evaluation of unlabeled pretraining for electrocardiogram foundation model performance. One-dimensional vision transformer (1D-ViT) masked autoencoders were pretrained across increasing ECG volumes and fine-tuned for rhythm, morphology, diagnostic, and structural heart disease tasks. Models pretrained at ≤400,000 ECGs failed to consistently exceed controls without self-supervised pre-training, whereas 600,000–800,000 ECGs improved AUROC across tasks, suggesting a minimum threshold for effective ECG representation learning.

## Main Text

The 12-lead electrocardiogram (ECG) is the most widely acquired cardiovascular test and contains embedded information about cardiac structure, function, and conduction. In addition to apparent diagnoses such as arrhythmia, deep learning models can identify substrate for future arrhythmia, ventricular dysfunction, and structural heart disease from ECG waveforms, implying the ECG encodes latent physiologic signatures not readily apparent to human readers [1–3]. However, most ECG artificial intelligence models remain task-specific, limiting their adaptability across clinical settings and disease categories, and have generally required large numbers of labeled datasets for acceptable performance.

Foundation models offer a different paradigm, starting with broad pretraining, often by self-supervision with unlabelled data, followed by supervised fine-tuning for multiple downstream tasks [4]. For both language and vision applications, performance often improves with data, model size, and compute, motivating empirical scaling laws [5,6]. Whether such similar scale-dependent behavior applies to ECG foundation models remains unclear.

Recent ECG foundation models have used masked reconstruction, contrastive learning, spatiotemporal masking, and multimodal representation learning to improve label efficiency and generalization [7–11]. We focused on masked autoencoding, in which a model learns to reconstruct missing portions of the input signal from visible waveform segments [12]. Although these approaches can improve downstream performance and label efficiency, the amount of unlabeled ECG data required for useful transfer is unknown. This matters because well-curated ECG repositories are not equally accessible and small institutional datasets may be insufficient for effective pretraining.

We evaluated whether ECG foundation model performance improves with increasing unlabeled pretraining scale. Using MIMIC-IV-ECG, we pretrained a one-dimensional vision transformer masked autoencoder on progressively larger subsets of unlabeled ECGs, ranging from 100,000 to 800,000 waveforms, and compared each pretrained model with a non-pretrained control (Fig 1.) [13]. Downstream evaluation was performed across clinically distinct task families using PTB-XL rhythm, PTB-XL form, PTB-XL diagnostic labels, and EchoNext structural heart disease labels [3,14]. All models used the same architecture and fine-tuning strategy, allowing the effect of pretraining scale to be isolated.

**Figure 1.**
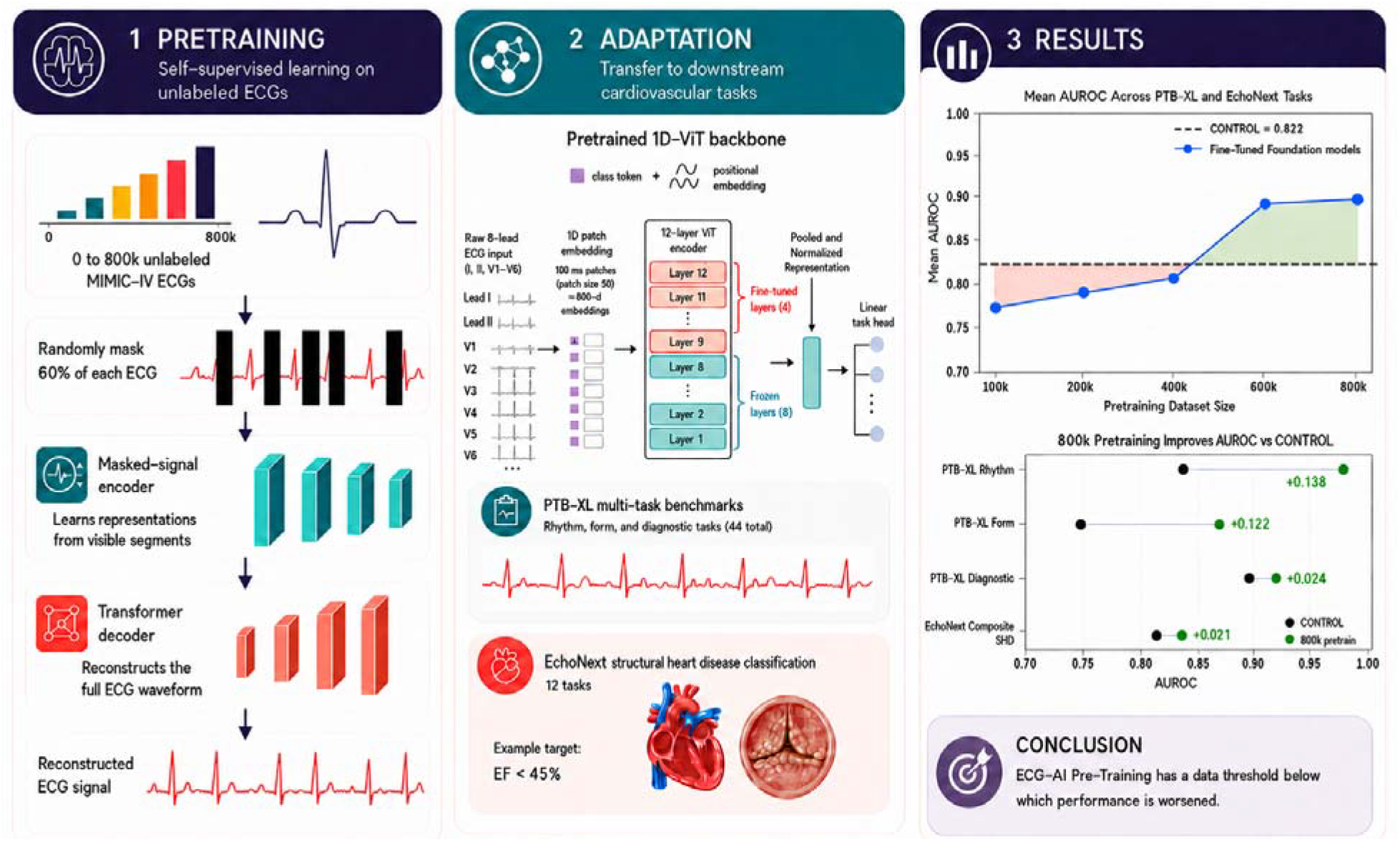
Study design for evaluating pretraining scale in ECG foundation models. A one-dimensional vision transformer was pretrained using a masked autoencoder objective across increasing subsets of unlabeled MIMIC-IV-ECG waveforms, ranging from 0 to 800,000 ECGs. During pretraining, 60% of the ECG signal was randomly masked and the model was trained to reconstruct the original waveform. The pretrained encoder was then fine-tuned for downstream ECG interpretation tasks using PTB-XL rhythm, form, and diagnostic labels and EchoNext structural heart disease labels. During fine-tuning, the classification head and top four transformer blocks were updated, while the lower eight transformer blocks were frozen. The study tested whether increasing unlabeled ECG pretraining scale improves downstream AUROC and whether a minimum pretraining scale threshold is required for effective transfer.

Pretraining scale demonstrated a clear threshold effect (Fig 2). Models pretrained on smaller subsets did not consistently outperform the non-pretrained control. In contrast, models pretrained on 600,000 and 800,000 ECGs improved performance across all evaluated task families. The largest gains occurred for PTB-XL rhythm and form labels, with the 800,000-ECG model improving mean AUROC by 0.138 for rhythm tasks and 0.122 for form tasks compared with control. Smaller but consistent improvements were observed for PTB-XL diagnostic labels and EchoNext structural heart disease labels, with mean AUROC gains of 0.024 and 0.021, respectively.

**Figure 2.**
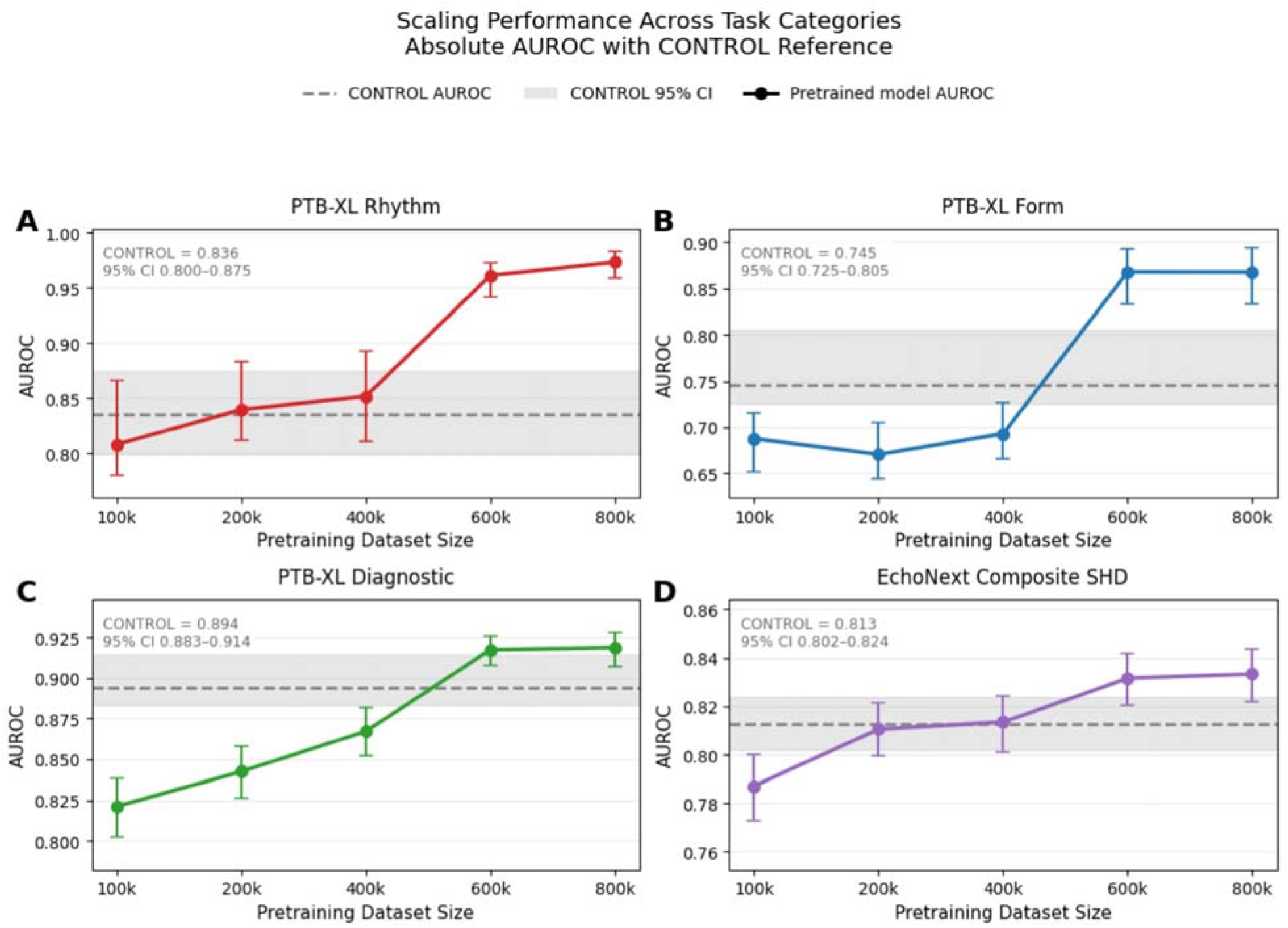
Downstream AUROC performance across pretraining scale and task families. AUROC performance is shown across increasing ECG pretraining dataset sizes for four downstream task families: a) PTB-XL rhythm labels, b) PTB-XL form labels, c) PTB-XL diagnostic labels, and d) EchoNext composite structural heart disease labels. Dashed horizontal lines indicate the non-pretrained control AUROC, and shaded regions indicate the 95% confidence interval for the control model. Models pretrained at smaller scales did not consistently exceed the non-pretrained control, whereas models pretrained on 600,000 and 800,000 ECGs improved performance across all task families. At 800,000 ECGs, mean AUROC improved by 0.138 for PTB-XL rhythm, 0.122 for PTB-XL form, 0.024 for PTB-XL diagnostic, and 0.021 for EchoNext composite structural heart disease tasks.

This pattern is clinically and methodologically plausible. Rhythm and form labels are closely related to waveform features directly present in the ECG, including periodicity, conduction intervals, QRS morphology, repolarization, and lead-specific activation patterns. These features align naturally with a masked waveform reconstruction objective. Diagnostic and structural heart disease labels may be more indirectly encoded in the surface ECG, reflecting subtler physiologic signatures rather than visually obvious morphology. The smaller absolute gains for these tasks therefore likely reflect greater label complexity and weaker direct signal-to-label correspondence. Nevertheless, improvement across structural heart disease tasks suggests that sufficiently large-scale pretraining can learn representations extending beyond narrow rhythm or morphology classification.

Structural heart disease performance varied across EchoNext labels but generally improved with increasing pretraining scale (Fig 3.). The 600,000- and 800,000-ECG models exceeded the non-pretrained control for most structural heart disease outcomes and outperformed the published EchoNext-mini baseline for all but one label [3]. For most tasks, performance approached the larger EchoNext model despite using substantially less labeled data. These findings support the possibility that unlabeled ECG pretraining can improve label efficiency for cardiovascular phenotyping, particularly when downstream labels require echocardiographic linkage or other resource-intensive annotation.

**Figure 3.**
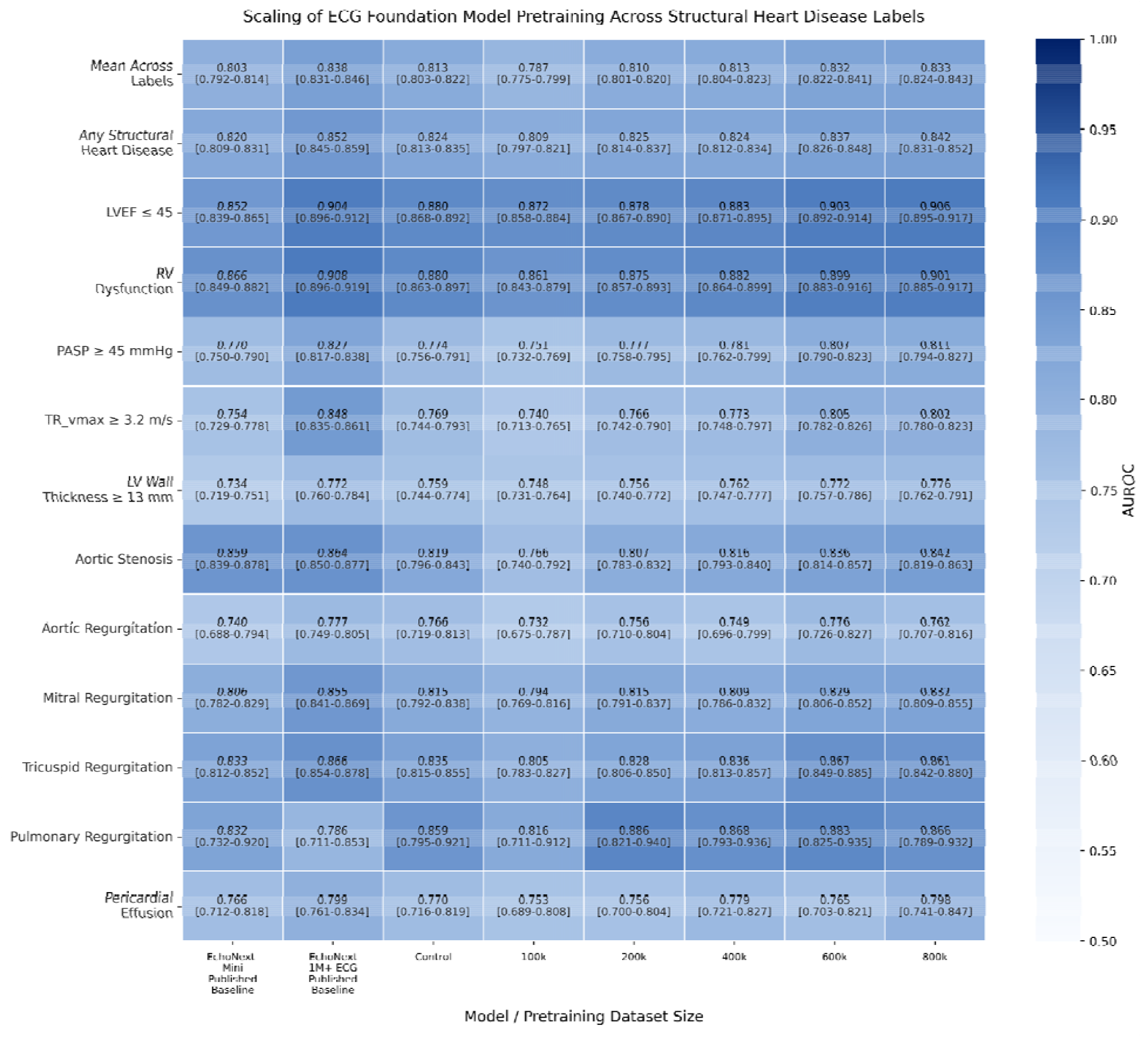
Task-level EchoNext structural heart disease performance across pretraining scale. Heatmap showing AUROC performance across individual EchoNext structural heart disease labels for EchoNext-Mini, EchoNext, the non-pretrained control, and models pretrained on increasing ECG dataset sizes from 100,000 to 800,000 ECGs. Larger pretraining scales generally improved performance across structural heart disease labels, although gains varied by task. The 600,000- and 800,000-ECG models exceeded the non-pretrained control for most labels and approached published EchoNext performance for several tasks, supporting the concept that large-scale unlabeled ECG pretraining may improve label efficiency for structural heart disease prediction.

The results have practical implications for ECG foundation model development. Self-supervised pretraining is often assumed to improve downstream performance regardless of scale, but our findings suggest that sub-scale pretraining is insufficient and may be counter-productive. The amount of unlabeled data used for pretraining appears to be a central design variable. At small scales, pretraining may not expose the model to enough variation in rhythm, morphology, acquisition quality, patient characteristics, and disease states to learn robust transferable representations. In that setting, the pretrained initialization may provide little advantage over supervised training from scratch. Larger pretraining sets may better capture the diversity of ECG phenotypes required for generalizable transfer.

These findings should be interpreted as evidence of a pretraining scale relationship rather than a definitive ECG scaling law. Formal scaling laws require evaluation across broader ranges of data, model size, and compute, often spanning orders of magnitude and permitting quantitative curve fitting [5,6]. In this study, we varied pretraining data volume while holding model architecture, masking objective, and fine-tuning strategy fixed. Thus, the central conclusion is not that ECG foundation models follow a universal power-law relationship, but that downstream transfer appears scale-dependent and may require a minimum amount of unlabeled ECG data before benefits emerge.

Several limitations should be considered. First, the maximum pretraining scale was 800,000 ECGs, so larger corpora are needed to determine whether performance continues to improve or saturates. Second, we evaluated a single architecture and masked autoencoder objective; different model sizes, masking ratios, reconstruction targets, contrastive objectives, or multimodal approaches may shift the observed threshold [8–12]. Third, downstream fine-tuning updated only the classification head and upper transformer blocks; full fine-tuning, linear probing, or parameter-efficient adaptation may produce different scale-performance relationships. Fourth, although we used established public benchmarks, additional validation across institutions, acquisition systems, populations, and clinical settings is needed to assess generalizability [3,13,14]. Finally, comparisons with published structural heart disease baselines should be interpreted cautiously when preprocessing, label definitions, training data, or evaluation protocols differ.

In summary, ECG foundation model pretraining demonstrated a scale-dependent threshold effect. Smaller-scale pretraining failed to consistently improve downstream transfer, whereas larger-scale pretraining improved performance across rhythm, form, diagnostic, and structural heart disease tasks. These findings suggest that effective ECG foundation model development may require substantial unlabeled data and that pretraining scale should be treated as a central methodological consideration.

## Methods

### Study design

We performed a pretraining scale analysis to evaluate whether self-supervised ECG foundation model performance improves with increasing unlabeled data volume. A one-dimensional vision transformer was pretrained using a masked autoencoder objective on progressively larger subsets of MIMIC-IV-ECG and subsequently fine-tuned on independent downstream benchmarks. Pretraining scale was the primary experimental variable. Model architecture, ECG preprocessing, masking ratio, and downstream fine-tuning procedures were otherwise held constant across pretrained models. A non-pretrained control model with the same architecture was trained from randomly initialized weights on each downstream task (Figure 1.).

### Pretraining dataset

Unlabeled ECG waveforms were drawn from MIMIC-IV-ECG [13]. We constructed pretraining subsets of increasing size: 100,000, 200,000, 400,000, 600,000, and 800,000 ECGs. Subsets were sampled from the available MIMIC-IV-ECG pretraining pool without using downstream task labels. Each subset was used to train a separate masked autoencoder model. The 800,000-ECG model represented the largest pretraining scale evaluated.

### Downstream datasets and labels

Downstream evaluation used PTB-XL and EchoNext Mini [3,14]. PTB-XL was used to evaluate three task families: rhythm labels, form labels, and diagnostic labels. EchoNext Mini was used to evaluate structural heart disease labels, including left ventricular systolic dysfunction and valvular abnormalities. Predefined dataset folds were used for training, validation, and testing when available. For all downstream tasks, model selection was performed on validation performance and final performance was reported on held-out test data.

### ECG preprocessing

All ECGs were represented as fixed-length waveform tensors. Signals were formatted as eight-lead ECGs consisting of leads I, II, and V1–V6. The remaining limb leads were omitted because they are linearly derivable from leads I and II. Each ECG was represented as an 8 × 5000 tensor, corresponding to eight leads over a standardized 10-second recording at 500 Hz. EchoNext waveforms originally sampled at 250 Hz were resampled to 500 Hz before model input. ECGs were normalized using dataset-specific mean and standard deviation values computed from the training data.

### Model architecture

We used a one-dimensional vision transformer architecture adapted for ECG waveform modeling. Each 10-second ECG was divided into non-overlapping waveform patches of 50 samples, corresponding to 100 milliseconds at 500 Hz. Each patch was flattened and linearly projected into an embedding space of dimension 800. A trainable class token was prepended to the patch sequence, and trainable positional embeddings were added to preserve temporal order.

The transformer encoder consisted of 12 encoder blocks. Each block used pre-layer normalization, multi-head self-attention with 8 attention heads, residual connections, and a multilayer perceptron with GELU activation and hidden dimension 3200. During downstream classification, the encoded patch representations were aggregated using global average pooling, followed by layer normalization and a linear task-specific classification head. For multilabel tasks, the output layer produced one logit per label.

### Masked autoencoder pretraining

Self-supervised pretraining used a masked autoencoder objective. Each 10-second, 8-lead ECG was divided into fixed-length waveform patches of 50 samples, corresponding to 100 ms at a sampling frequency of 500 Hz. During pretraining, 60% of patches were randomly masked, and the model was trained to reconstruct the original waveform values from the remaining visible patches. This reconstruction objective encouraged the encoder to learn representations of local ECG morphology, temporal continuity, rhythm structure, and interlead relationships without requiring clinical labels.

A separate model was pretrained for each MIMIC-IV-ECG subset size. Pretraining used a base masked time-series transformer architecture with the same input representation, masking ratio, preprocessing pipeline, and optimization strategy across all pretraining scales. Models were pretrained using AdamW optimization with a learning rate of 1 × 10□^3^, cosine learning rate scheduling, a batch size of 384, binary cross-entropy loss with smoothing of 0.01, and up to 300 epochs. No data augmentation was applied, and pretraining was performed on a single GPU. This fixed experimental design ensured that differences in downstream performance reflected pretraining data volume rather than changes in architecture or training configuration.

### Downstream fine-tuning

For each pretrained model, downstream fine-tuning was performed separately for each task family. The classification head and top four transformer encoder blocks were updated during fine-tuning, while the lower eight transformer blocks were frozen. This partial fine-tuning strategy was chosen to preserve general pretrained representations while allowing task-specific adaptation in the upper layers. The non-pretrained control used the same architecture and downstream training pipeline but was initialized without pretrained weights.

Models were trained for multilabel classification using binary cross-entropy loss with label smoothing of 0.001. Fine-tuning used AdamW optimization with a learning rate of 5 × 10□□, no weight decay, a batch size of 256, and cosine learning rate scheduling. Models were trained for up to 100 epochs, with early stopping based on validation AUROC using a patience of 10 epochs. Fine-tuning was performed on a single GPU with a fixed random seed to support reproducibility.

### Performance evaluation

The primary performance metric was area under the receiver operating characteristic curve. AUROC was computed for each label independently. Mean AUROC was then calculated within each task family: PTB-XL rhythm, PTB-XL form, PTB-XL diagnostic, and EchoNext structural heart disease. For EchoNext, both aggregate structural heart disease performance and individual label-level results were evaluated.

Performance was compared across pretraining scales and against the non-pretrained control. Absolute AUROC differences were calculated by subtracting control performance from pretrained model performance for each task family. Published EchoNext-mini and EchoNext model results were used as contextual baselines for structural heart disease tasks when comparable labels were available [3].

## Statistical analysis

Uncertainty was estimated using nonparametric bootstrapping with 2000 repetitions. For each bootstrap replicate, the test set was resampled with replacement, performance metrics were recomputed, and empirical 95% confidence intervals were derived from the bootstrap distribution. The same resampling framework was applied across task families and structural heart disease labels. Results were summarized as AUROC values with 95% confidence intervals and as absolute AUROC differences relative to the non-pretrained control.

### Computational environment

All model pretraining and downstream fine-tuning experiments were performed on a single GPU. This design was intended to reflect a computationally constrained but feasible training setting for institutional ECG foundation model development. The architecture and training strategy were therefore selected to balance representation capacity with practical hardware requirements.

### Role of pretraining scale

The primary analysis evaluated whether downstream performance improved monotonically or threshold-dependently with increasing pretraining data volume. Because model size, architecture, masking ratio, downstream datasets, fine-tuning strategy, and evaluation procedures were fixed, observed differences across models were attributed primarily to the scale of unlabeled ECG pretraining. This design allowed assessment of whether small-to-moderate pretraining sets were sufficient for useful transfer or whether larger unlabeled ECG corpora were required before consistent downstream gains emerged.

## Data Availability

All ECG data mentioned (MIMIC-IV, EchoNext, and PTB-XL) is accessible through physionet.

## Code Availability

The code to pretrain this model with MIMIC-IV and fine tune this model for PTB-XL are available on this github page: https://github.com/4dm-labs/ecgflow. The EchoNext data loader and fine-tune script will be available in this repository at the time of publication.

## Acknowledgments

VLM and SNG are supported by the National Institutes of Health and Wellcome Leap Visible. The funding sources had no role in the study design, data collection, analysis, interpretation, manuscript writing, or decision to submit the manuscript for publication.

## Author contributions

RS, VLM and IN conceptualized and established the study design, and wrote the manuscript draft BF and RS performed statistical analysis ZL, SY, SNG, and ES contributed to study evaluation, data review, and manuscript writing VLM and IN supervised the study and contributed to design and writing

